# Multiomic characterisation of high grade serous ovarian carcinoma enables high resolution patient stratification

**DOI:** 10.1101/2022.01.07.22268840

**Authors:** Robert L Hollis, Alison M Meynert, Caroline O Michie, Tzyvia Rye, Michael Churchman, Amelia Hallas-Potts, Ian Croy, W. Glenn McCluggage, Alistair R W Williams, Clare Bartos, Yasushi Iida, Aikou Okamoto, Brian Dougherty, J. Carl Barrett, Ruth March, Athena Matakidou, Patricia Roxburgh, Colin A Semple, D Paul Harkin, Richard Kennedy, C Simon Herrington, Charlie Gourley

**Affiliations:** Nicola Murray Centre for Ovarian Cancer Research, Cancer Research UK Edinburgh Centre, MRC Institute of Genetics and Cancer, University of Edinburgh, UK; MRC Human Genetics Unit, MRC Institute of Genetics and Cancer, University of Edinburgh, UK; Edinburgh Cancer Centre, Western General Hospital, NHS Lothian, UK; Department of Pathology, Belfast Health and Social Care Trust, Belfast, UK; Division of Pathology, The Royal Infirmary of Edinburgh, Edinburgh, UK; The Jikei University School of Medicine, Tokyo, Japan; Translational Medicine, Oncology R&D, AstraZeneca, Waltham, MA, USA; Precision Medicine and Biosamples, Oncology R&D, AstraZeneca, Cambridge, UK; Centre for Genomics Research, Discovery Sciences, BioPharmaceuticals R&D, AstraZeneca, Cambridge, UK; Institute of Cancer Sciences, Wolfson Wohl Cancer Research Centre, University of Glasgow, UK; Beatson West of Scotland Cancer Centre, Glasgow, UK; Almac Diagnostics, Craigavon, UK; Centre for Cancer Research and Cell Biology, Queen’s University Belfast, UK

**Keywords:** high grade serous ovarian cancer, genomics, transcriptomics, survival, treatment response

## Abstract

**Background:** High grade serous ovarian carcinoma (HGSOC) is the most common type of ovarian cancer; most patients experience disease recurrence which accumulates chemoresistance, leading to treatment failure. Previous investigations have characterised HGSOC at the genomic and transcriptomic level, identifying subtypes of patients with differential outcome and treatment response. However, the relationship between molecular events identified at the gene sequence, gene copy number and gene expression levels remains poorly defined.

**Methods:** We perform multi-layer molecular characterisation of a large retrospective HGSOC cohort (n=362) with detailed clinical annotation to interrogate the relationship between patient groups defined by gene mutation, copy number events, gene expression patterns and infiltrating immune cell burden. We construct a high resolution picture of the molecular landscape in HGSOC and identify features of tumours associated with distinct clinical behaviour in patients.

**Results:** *BRCA2*-mutant (*BRCA2*m) and *EMSY*-overexpressing cases demonstrated prolonged survival (multivariable hazard ratio 0.40 and 0.53) and higher chemotherapy response rates at first- and second-line treatment. *CCNE1*-gained (*CCNE1*g) cases demonstrated shorter survival (multivariable hazard ratio 1.52, 95% CI 1.10-2.10), under-representation of FIGO stage IV cases (P=0.017) and no significant difference in treatment response. We demonstrate marked overlap between the TCGA- and derived subtypes: the TCGA DIF, IMR, PRO and MES subtypes correlated with the Tothill C4, C2, C5 and C1 subtypes (P<0.001). IMR/C2 cases displayed higher *BRCA1/2*m frequency (25.5% and 32.5%) and significantly greater infiltration of immune cells (P<0.001), while PRO/C5 cases had the highest *CCNE1*g rate (23.9% and 22.2%) and were uniformly low in immune cell infiltration. The survival benefit for cases with aberrations in homologous recombination repair (HRR) genes was apparent across all transcriptomic subtypes (hazard ratio range 0.48-0.68). There was significant co-occurrence of RB loss and HRR gene aberrations (P=0.005); RB loss was further associated with favourable survival within cases harbouring HRR aberrations (multivariable hazard ratio 0.50, 95% CI 0.30-0.84).

**Conclusions:** These data paint a high resolution picture of the molecular landscape in HGSOC, better defining patients who may benefit most from specific molecular therapeutics and highlighting those for whom novel treatment strategies are needed to improve outcomes.

## 1. BACKGROUND

High grade serous ovarian carcinoma (HGSOC) is the most common form of tubo-ovarian cancer. The majority of HGSOC patients are diagnosed at advanced stage and experience poor prognosis, with a five-year survival of approximately 30% in this population [1]. While the majority of HGSOC demonstrate high levels of intrinsic sensitivity to platinum-based chemotherapy, most patients experience disease recurrence which accumulates therapy resistance, leading to progressively shorter treatment-free intervals until patients eventually succumb to disease [2, 3].

In the hope of identifying therapeutically exploitable disease biology, a wealth of data have been produced over the last two decades characterising the genomic and transcriptomic landscape of HGSOC [4-7]. At the gene sequence level, identification of mutational disruption in *BRCA1* and *BRCA2* (*BRCA1*/*2*m) has ultimately paved the way for integration of poly-(ADP-ribose) polymerase (PARP) inhibitor use into routine care for some patients [8-10]. Indeed, there continues to be an intense research effort surrounding mechanisms and implications of homologous recombination DNA repair (HRR) disruption beyond *BRCA1*/*2*m [11]; these include mutation of non-*BRCA1*/*2* HRR genes [12], large-scale genomic variants disrupting *BRCA1*/*2* [13], epigenetic inactivation of HRR players such as *BRCA1* and *RAD51C* [5, 14], and overexpression of the BRCA2 regulator EMSY [15, 16].

At the gene expression level, numerous studies have characterised HGSOC samples, endeavouring to identify clinically meaningful transcriptomic subtypes of disease or expression signatures predictive of survival risk [4, 5, 7, 17, 18]. Most notably, Tothill et al. [4] and the TCGA investigators [5] each identified multiple transcriptomic subtypes, associating these with differential survival profiles. These analyses have identified favourable outcome in patients with tumours harbouring expression profiles suggestive of active immune engagement (TCGA IMR subtype [5, 19], Tothill C2 subtype [4]) – consistent with earlier reports of favourable outcome in cases with high levels of cytotoxic T cell infiltration [20, 21]. However, transcriptomic subtyping is not currently used for clinical prognostication or stratification of HGSOC patients, despite some investigators reporting differential sensitivity of these groups to agents such as bevacizumab [22].

While multiple investigators have characterised either the genomic or transcriptomic landscape of HGSOC, few have investigated the relationship between genomic and transcriptomic features. Moreover, the relationship between these events and recently identified recurrent disruption of RB and PTEN in HGSOC is poorly understood [6]. Integration of multiple layers of molecular characterisation is required to paint a granular picture of the molecular landscape in HGSOC to better inform rationally designed trials of novel treatment regimens or combination therapy strategies. Indeed, some investigators have suggested that transcriptomic subtypes of HGSOC that appear to derive greatest benefit from anti-angiogenic agents may be depleted for *BRCA1*/*2*m cases who benefit most from PARP inhibition [5, 22]. This notion is consistent with mixed results observed from the addition of anti-angiogenic agents to PARP inhibitors (PARPi) dependent on patient selection, therapy line and agent combinations [23], exemplifying the need for comprehensive multi-layer molecular characterisation to inform patient selection for future investigations of novel treatment approaches.

Here we perform matched genomic and transcriptomic characterisation of a large, well annotated HGSOC cohort, dissecting the relationship between patient groups defined at the gene sequence, gene copy number and gene expression level.

## 2. METHODS

### 2.1 Patient cohort

539 ovarian cancer patients treated at the Edinburgh Cancer Centre met the following study inclusion criteria: primary ovarian, peritoneal or fallopian tube carcinoma (any histological type) diagnosed prior to 2007; available formalin-fixed paraffin-embedded (FFPE) treatment-naïve surgical specimen; first-line platinum-containing chemotherapy; minimum 3-year follow-up. Pathology review of H&E-stained slides was undertaken by expert gynaecological pathologists (WGM, ARWW, CSH) to identify HGSOC cases (figure S1); immunohistochemistry (IHC) for p53 and WT1 was used to clarify cases of uncertain histological type (HGSOC: WT1 positive, p53 aberrant mutation-type expression pattern) (Supplementary Methods Section 1) (figure S1). Ethical approval was obtained from South East Scotland Human Annotated Bioresource (Lothian NRS Bioresource Ethics Committee reference 15/ES/0094-SR705 and SR752). The need for consent was waived by the ethics committee due to the retrospective nature of the study.

### 2.2 Genomic characterisation

H&E stained slides were marked to identify tumour areas of high cellularity and used as a guide for macrodissection of 10µm FFPE sections for DNA extraction. DNA extraction was performed using the QIAamp FFPE DNA Kit and Qiagen Deparaffinisation Solution (Qiagen). Extracted DNA was quantified by high sensitivity Qubit assay. *CCNE1* and *EMSY* copy number (CN) were quantified by TaqMan qPCR (Supplementary Methods Section 2).

High throughput sequencing was performed using a custom Integrated DNA Technologies (IDT) gene capture panel with unique molecular indices (UMIs) (Supplementary Methods Section 3). Whole genome libraries were generated, pooled for target capture and sequenced using an Illumina NextSeq 550 at the Edinburgh Clinical Research Facility, Western General Hospital, Edinburgh, UK. The median per-sample mean target coverage was 593X (range 205-3278X). Reads were processed using the bcbio v1.0.6 high throughput sequence analysis pipeline (Supplementary Methods Section 4). Consensus reads aligned to hg38 underwent variant calling using a majority vote system from three variant callers (Freebayes, VarDict and Mutect2). Called variants were annotated using the Ensembl VEP v90.9 against Ensembl release 90 and filtered to retain only functional variation (Supplementary Methods Section 5).

### 2.3 Transcriptomic subtyping

Transcriptomic data for the cohort were available from previous work identifying transcriptomic subtypes of HGSOC [15, 17], including *EMSY* overexpression status (Supplementary Methods Section 6). TCGA (MES, PRO, IMR, DIF) and Tothill (C1, C2, C4, C5) transcriptomic subtyping calls were made with the consensusOv R package using the consesusOv and Helland approaches [19] (Supplementary Methods Section 6).

### 2.4 Immune cell infiltration analysis

Tumour infiltrating CD3-positive and CD8-positive immune cells were quantified by IHC of constructed tumour tissue microarrays (TMAs) (Supplementary Methods Section 7); marker-positive cell burden was quantified as percentage positive cells within tumour areas using QuPath version 0.1.2 [24].

### 2.5 Detection of PTEN and RB loss by immunohistochemistry

PTEN and RB protein loss was detected by IHC using sections of the HGSOC TMA (Supplementary Methods Section 8). Loss was defined as complete absence of positive staining in tumour cells with confirmed corresponding positive internal control stromal staining.

### 2.6 Copy number analysis from off-target sequencing reads

Copy number analysis was performed using CopywriteR [25] (Supplementary Methods Section 9): off target reads were used to estimate the relative copy number of 50kB genome segments across each chromosome, using the alignment bam files from the above sequencing analysis workflow. For quantification of CN alteration burden, adjacent 50kB segments of gain/loss representing the same large CN event were merged prior to quantification (Supplementary Methods Section 9).

### 2.7 Clinical annotation

Baseline clinicopathological features and outcome data were extracted from the Edinburgh Ovarian Cancer Database [26], alongside chemotherapy response data (Supplementary Methods Section 10). Overall survival (OS) and progression-free survival (PFS) were defined as the time from pathologically confirmed diagnosis to patient death and disease progression or recurrence, respectively (Supplementary Methods Section 9).

### 2.8 Statistical analyses

All statistical analyses were performed using R version 4.0.3 (R Foundation for Statistical Computing, Vienna, Austria). Comparisons of frequency were performed using the Chi-squared test or Fisher’s exact test, as appropriate. Between-group comparisons of continuous variables was performed using the Mann Whitney-U test. Survival analysis was performed using Cox proportional hazards regression models and reported as hazard ratios (HR) with 95% confidence intervals (95% CI). For survival analysis adjusted for other clinicopathological factors, multivariable hazard ratios (mHR) are reported. Median follow-up time was calculated by the reverse Kaplan-Meir method. Adjustment for multiple testing was applied using the Bonferroni method.

## 3. RESULTS

### 3.1 Cohort characteristics

Of 539 ovarian cancer cases that met eligibility criteria, 362 were classified as HGSOC following pathology review and underwent molecular characterisation (n=27 insufficient tumor, n=131 non-HGSOC, n=1 failed sequencing library preparation, n=8 failed quality control) (figure S1). Clinicopathological features of the study cohort are summarised in table 1. The median follow-up time was 15.0 years.

**Table 1.** Clinicopathological features of high grade serous carcinoma

|  |  | N | % |
| --- | --- | --- | --- |
| **Total cases** | N | 362 |  |
| **Age at diagnosis** | Median years | 61 | Range 33-86 |
| **FIGO stage at diagnosis** | I | 15 | 4.3 |
|  | II | 31 | 8.8 |
|  | III | 237 | 67.5 |
|  | IV | 68 | 19.4 |
|  | NA | 11 | - |
| **RD following surgical debulking** | No visible RD (0cm) | 65 | 19.4 |
|  | Macro RD (0.1-2cm) | 66 | 19.7 |
| | Gross macro RD ($\geq 2$cm) | 187 | 55.8 |
|  | Macro RD of unknown size | 17 | 5.1 |
|  | Unknown | 27 | - |
| **First line chemotherapy** | Single-agent platinum | 217 | 59.9 |
|  | Platinum-taxane combination | 135 | 37.3 |
|  | Other platinum-containing regimes | 10 | 2.8 |
| **Vital status at last follow-up** | Alive | 36 | 9.9 |
|  | Deceased - died of OC | 307 | 84.8 |
|  | Deceased - other causes | 19 | 5.2 |
| **Median follow-up time** | Years | 15.0 (95% CI 12.8-19.9) |  |
| **Median PFS** | Years | 1.17 (95% CI 1.09-1.28) |  |
| **Median OS** | Years | 2.60 (95% CI 2.40-3.07) |  |

FIGO, International Federation of Gynecology and Obstetrics; NA, not available; Macro, macroscopic;
RD, maximal residual disease diameter; OC, ovarian carcinoma

### 3.2 Molecular landscape of high grade serous ovarian carcinoma

The frequency of *TP53* mutation was 98.1% (355 of 362 cases) (figure 1, table S1). 12.7% and 6.6% of cases harboured *BRCA1*m and *BRCA2*m. Eight cases (2.2%) demonstrated mutation of other HRR genes (3 *BRIP1*, 2 *CHEK2*, 1 *RAD51C*, 1 *PALB2*, 1 concurrent *BAP1* and *NBN*). 14.9% of cases displayed CN gain of *CCNE1* (*CCNE1*g) and 6.6% demonstrated amplification of *EMSY*. Tumours demonstrating *EMSY* amplification were enriched for *EMSY* mRNA-overexpressing cases (P<0.001) (Supplementary table 2); however, *EMSY* CN was a poor predictor of *EMSY* overexpression status (positive predictive value 0.42, 95% CI 0.22-0.63; negative predictive value 0.88, 95% CI 0.84-0.91).

**Figure 1.**
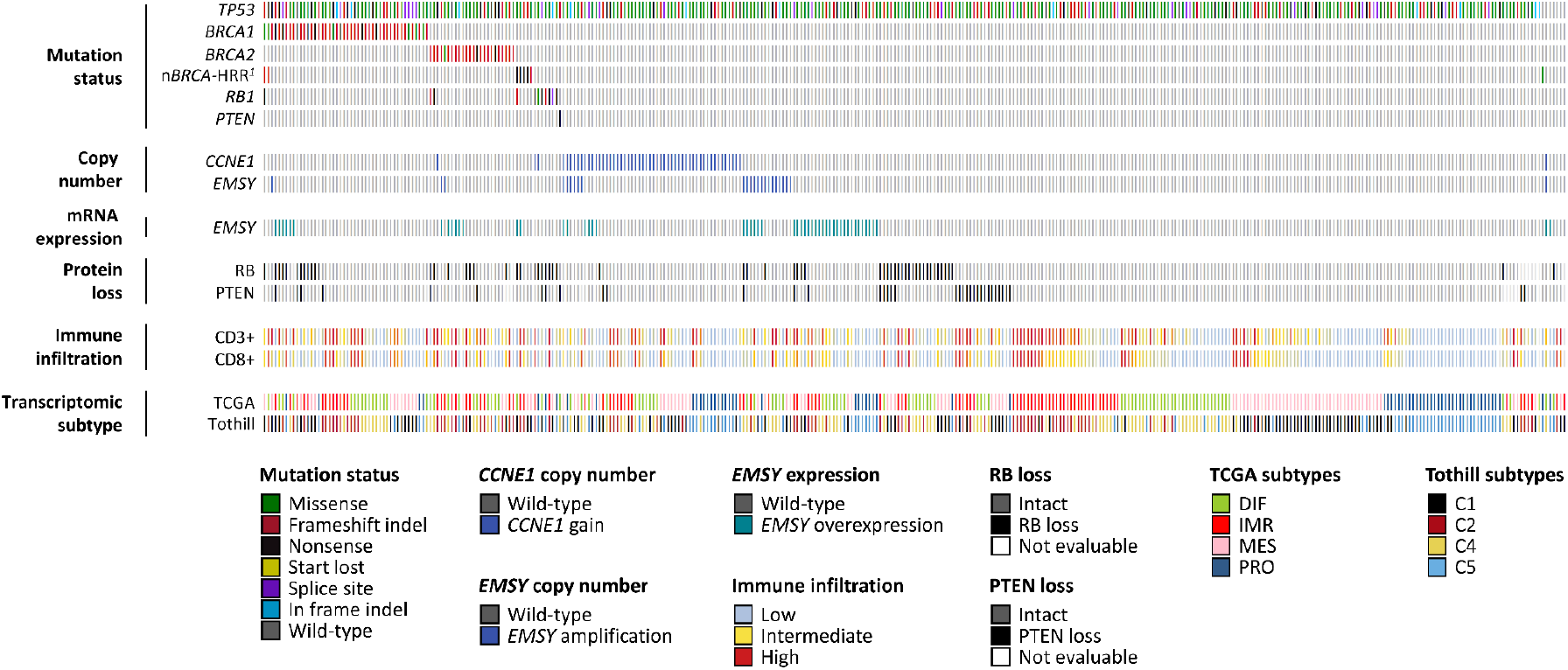
Molecular landscape of high grade serous ovarian carcinoma. ^1^Mutation in non-*BRCA1*/*2* homologous recombination repair (HRR) genes: 3 *BRIP1*, 2 *CHEK2*, 1 *RAD51C*, 1 *PALB2*, 1 concurrent *BAP1* and *NBN. CCNE1* copy number (CN) gain, ≥4 copies by Taqman CN assay. *EMSY* CN amplification, ≥6 copies by Taqman CN assay.

An HRR-centric stepwise taxonomy was constructed (figure 2A). Compared to the non-*CCNE1*g HRR-wild-type reference population (HRRwt, 55.8% of cases), *BRCA2*m and *EMSY*-overexpressing (8.6%) cases demonstrated favourable outcome (mHR for OS = 0.40, 95% CI 0.25-0.65 and 0.53, 95% CI 0.33-0.84) (figure 2A, figure S2). Stage IV cases were under-represented in the *CCNE1*g group (8.2% vs 23.4% in the HRRwt group, P=0.017); *CCNE1*g cases demonstrated significantly shorter survival after accounting for age, stage and debulking status (mHR for OS = 1.52, 95% CI 1.10-2.10). The *BRCA2*m and *EMSY*-overexpressing subgroups demonstrated the highest rates of complete response to first and second-line chemotherapy as determined by radiology or CA125 tumour marker (figure 2C). Complete response rate was higher in the *BRCA2*m, *EMSY*-overexpressing and *BRCA1*m cases compared to the HRRwt cases at first chemotherapy (P<0.001, P=0.009 and P=0.049 for complete GCIG CA125 response [confirmed normalisation from at least double upper limit of normal]); however, only *BRCA2*m and *EMSY*-overexpressing cases had a significantly higher complete response rate after adjusting for multiple testing (P-adj=0.001, P-adj=0.027, P-adj=0.148). At relapse, *BRCA2*m and *EMSY*-overexpressing cases retained a higher chemotherapy response rate (P=0.002 and P=0.037 for complete CA125 response, respectively). Chemotherapy response rate was similar in the *CCNE1*g and HRRwt groups at both primary treatment and relapse (figure 2C).

**Figure 2.**
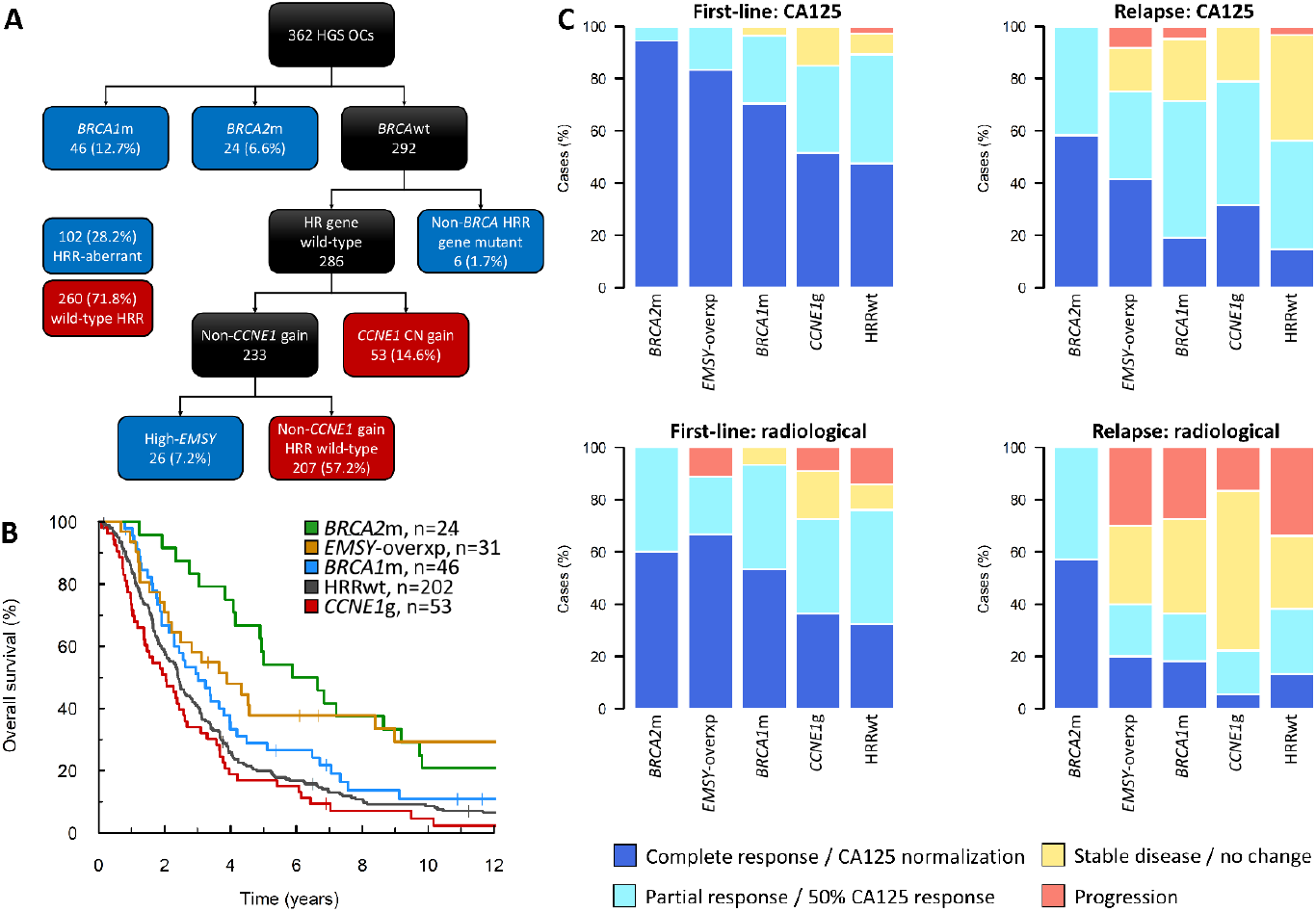
Homologous recombination repair pathway (HRR)-centric subtyping of high grade serous carcinoma. (A) HRR-centric classification taxonomy. (B) Overall survival profile of HRR-centric subtypes. (C) Chemosensitivity of HRR-centric subtypes at first-line treatment (left hand panels) and treatment for disease relapse (right hand panels) as determined by CA125 tumour marker (top panels) and radiology (bottom panels). *BRCA2*m, *BRCA2* mutant; *BRCA1*m, *BRCA1* mutant; *EMSY*-overxp; overexpression of *EMSY*; *CCNE1*g, gain of *CCNE1*; HRRwt, non-*CCNE1*g homologous recombination proficient.

### 3.3 Relationship between transcriptomic subtypes

Two transcriptomic subtyping approaches were used (TCGA subtypes: DIF, IMR, PRO, MES; Tothill subtypes: C1, C2, C4, C5). There was marked overlap between the subtyping approaches (P<0.0001) (figure 3A): PRO cases were overwhelmingly of the C5 subtype (91.0%, 61 of 67), while the vast majority of MES cases were of the C1 subtype (88.9%, 88 of 99). The DIF group comprised mainly C4 tumours (69.6%, 71 of 102), while IMR cases were mostly of the C2 subtype (66.0%, 62 of 94).

**Figure 3.**
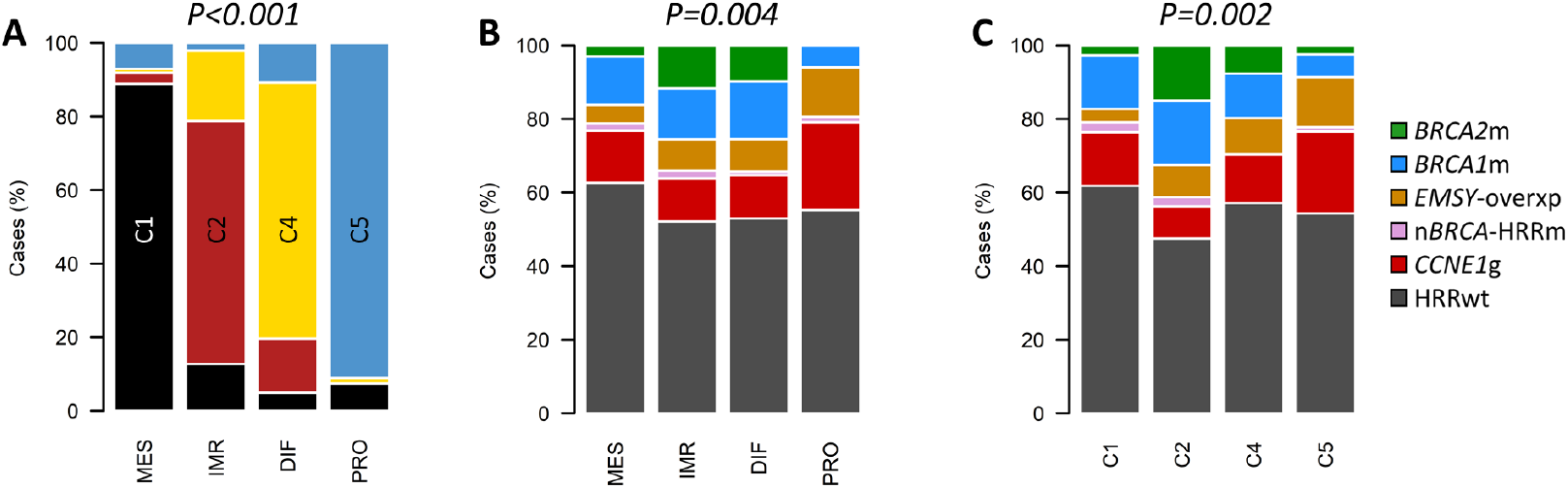
Relationship between subgroups methodologies. (A) Comparison of transcriptomic subgrouping approaches: composition of Tothill subtypes across each of the TCGA subtypes; labelled P value represents comparison of Tothill subtype frequency across all TCGA subtypes by Chi-squared test. (B) Distribution of homologous recombination repair (HRR)-centric subtypes across each of the TCGA transcriptomic subtypes; labelled P value represents comparison of *BRCA1*/*2*m frequency across all groups by Chi-squared test; P-adj=0.009. (C) Distribution of homologous recombination repair (HRR)-centric subtypes across each of the Tothill transcriptomic subtypes; labelled P value represents comparison of *BRCA1*/*2*m frequency across all groups by Chi-squared test; P-adj=0.003. *BRCA2*m, *BRCA2* mutant; *BRCA1*m, *BRCA1* mutant; *EMSY*-overxp; overexpression of *EMSY*; nBRCA-HRRm, non-*BRCA1*/*2* HRR gene mutation; *CCNE1*g, gain of *CCNE1*; HRRwt, non-*CCNE1*g homologous recombination proficient.

### 3.4 Genomic-transcriptomic correlates

There was marked association between HRR-centric and transcriptomic subtypes (figure 3B and 3C). Frequency of *BRCA1* and *BRCA2* mutation differed significantly between transcriptomic subtypes, with the highest and lowest *BRCA1*/*2*m rates in the IMR/C2 and PRO/C5 subtypes, respectively (25.5% and 32.5% vs 6.0% and 8.6%, P-adj=0.009 and 0.003) (figure 3B and 3C). Frequency of *CCNE1*g was highest in PRO/C5 tumours (23.9% in PRO, 22.2% in C5), while the C2 subtype demonstrated the lowest *CCNE1*g frequency (8.8%, P=0.002) (figure 3B and 4C).

Prolonged survival for HRR deficient cases was apparent across all transcriptomic subtypes (HR range 0.48-0.68) (figure S3 and S4). We did not observe any significant differences in overall burden of CN loss or gain events between transcriptional subtypes (figure S5).

### 3.5 Immune cell infiltration burden

The burden of tumour-infiltrating CD3+ and CD8+ cells was heterogeneous across samples, with higher infiltration associated with prolonged survival (figure S6). *BRCA2*m cases demonstrated the highest levels of CD3+ infiltration (figure 4A).

**Figure 4.**
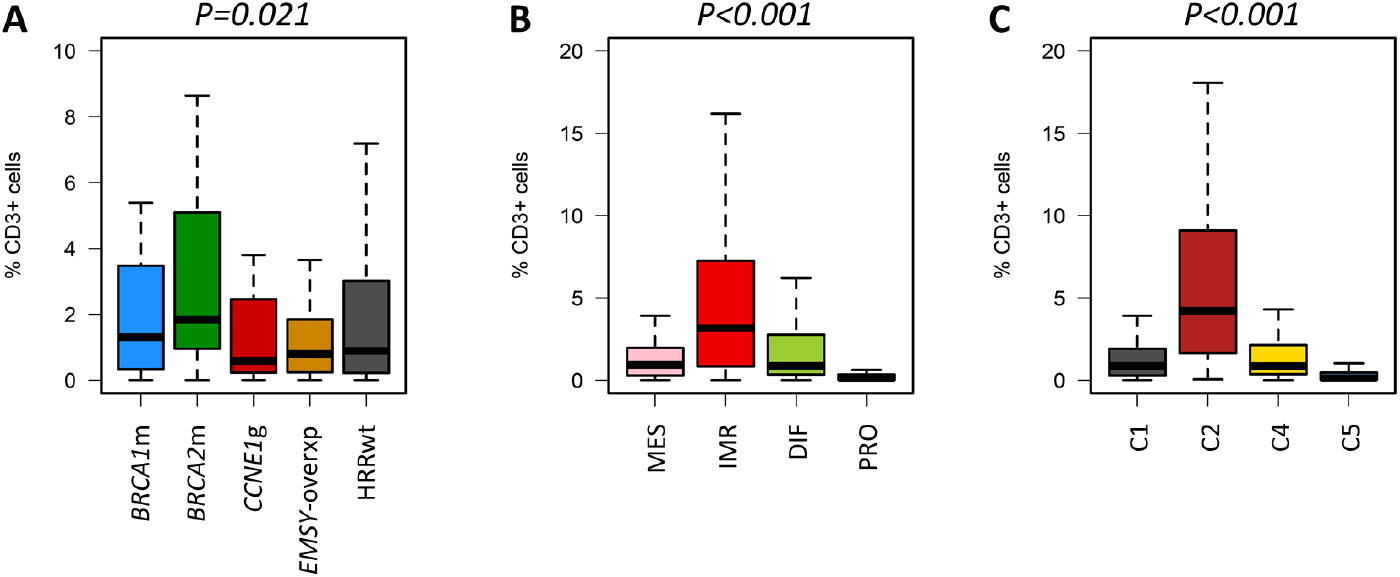
Tumour-infiltrating immune cells across high grade serous carcinoma subtypes. (A) CD3+ infiltration across HRR-centric subtypes; labelled P value represents comparison of *BRCA2*m and *CCNE1*g groups using the Mann Whitney-U test. (B) CD3+ infiltration across TCGA transcriptomic subtypes; labelled P value represents comparison of IMR and PRO groups using the Mann Whitney-U test. (C) CD3+ infiltration across Tothill transcriptomic subtypes; labelled P value represents comparison of C2 and C5 groups using the Mann Whitney-U test. *BRCA2*m, *BRCA2* mutant; *BRCA1*m, *BRCA1* mutant; *EMSY*-overxp; overexpression of *EMSY*; *CCNE1*g, gain of *CCNE1*; HRRwt, non-*CCNE1*g homologous recombination proficient.

Subtypes defined by both transcriptomic subgrouping methodologies demonstrated marked differences in infiltrating CD3+ (P-adj<0.0001) (figure 4B and 4C) and CD8+ cells (P-adj<0.0001) (figure S7). The IMR/C2 subtypes demonstrated the highest infiltration levels, while the PRO/C5 subtypes demonstrated uniformly low levels of infiltrating cells.

### 3.6 RB and PTEN loss in HGSOC

10.6% of cases (37 of 350 evaluable tumours) demonstrated PTEN protein loss (figure 1, figure 5A). PTEN loss was a rare event in tumours of the PRO/C5 subtypes (3.0% in PRO, 2.5% in C5) (figure S9A and S9B). Cases with loss of PTEN expression demonstrated significantly lower *PTEN* CN (P=0.0003) (figure S9A).

**Figure 5.**
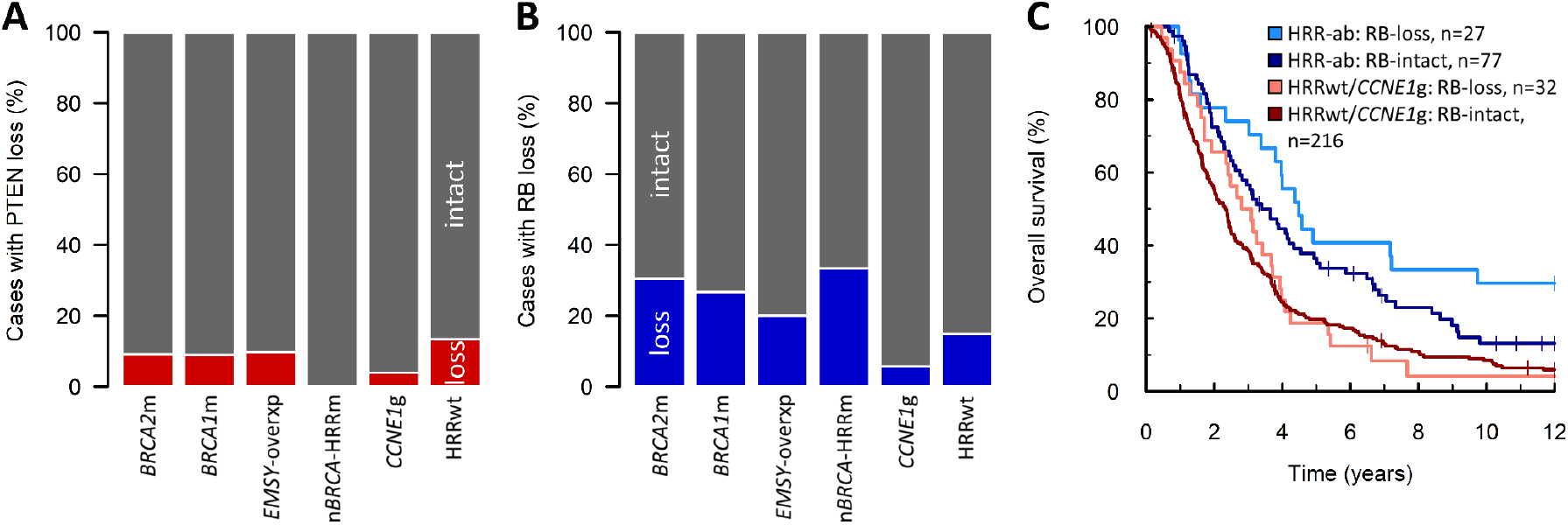
PTEN and RB loss in high grade serous ovarian carcinoma. (A) Frequency of loss of PTEN protein expression across homologous recombination repair (HRR)-centric subtypes. (B) Frequency of loss of RB protein expression across homologous recombination repair (HRR)-centric subtypes. (C) Impact of RB loss on survival in patients based on HRR status. Multivariable hazard ratio (mHR) for HRR-ab: RB-loss vs HRR-ab RB-intact=0.50, 95% CI 0.30-0.84; mHR for HRRwt/*CCNE1*g: RB-loss vs HRRwt/*CCNE1*g: RB-intact=0.71, 95% CI 0.48-1.06. *BRCA2*m, *BRCA2* mutant; *BRCA1*m, *BRCA1* mutant; *EMSY*-overxp; overexpression of *EMSY*; nBRCA-HRRm, non-*BRCA1*/*2* HRR gene mutation; *CCNE1*g, gain of *CCNE1*; HRRwt, non-*CCNE1*g HRR wild-type. HRR-ab, HRR-aberrant: *BRCA1*m, *BRCA2*m, *EMSY*-overxp or n*BRCA*-HRRm.

16.8% of cases (59 of 352 evaluable tumours) demonstrated loss of RB protein (figure 1, figure S8C and S8D). RB loss was ubiquitous among HGSOCs harbouring *RB1* mutation (11/11 cases demonstrating loss; P<0.001 vs 48/341 in the absence of *RB1* mutation) (figure 1). Cases demonstrating RB loss had a lower *RB1* CN (P=0.0258) (figure S9B) and there was significant co-occurrence between RB loss and PTEN loss (22.8% PTEN loss in RB-lost cases, 13/57 vs 7.7%, 22/285 evaluable cases; P=0.001) (figure 1).

RB loss was significantly enriched among cases with HRR gene aberrations (*BRCA1*m, *BRCA2*m, *EMSY*-overexpression or non-*BRCA*-HRR mutation) (26.0%, 27/104 evaluable cases vs 12.9%, 32/248; P=0.005) (figure 5A) and was a rare event among *CCNE1*g cases (5.7%). In cases with HRR gene aberrations, RB loss was associated with significantly longer survival (mHR for OS=0.50, 95% CI 0.30-0.84) (figure 5B); conversely, RB loss was not associated with significant differences in survival within the remaining population (mHR=0.71, 95% CI 0.53-1.06) (figure 5B).

## 4. DISCUSSION

Substantial advancements in our understanding of HGSOC biology have been made over the last two decades, with many studies characterising HGSOC cases at the gene sequence and gene expression level [4-6, 19]. These investigations have identified subgroups of patients with differential outcome and therapy sensitivity, paving the way for molecular stratification of HGSOC patient care [8, 22, 27-29]. However, the relationship between features described at the genomic and transcriptomic level is poorly understood. We present matched genomic-transcriptomic characterisation – alongside identification of other molecular features, including RB expression loss, PTEN expression loss, and immune cell infiltration – in a large pathologically-confirmed HGSOC cohort with detailed clinical annotation and extensive follow-up, revealing marked correlation across these levels of molecular characterisation.

We utilised two transcriptomic subtyping approaches within our dataset: there was substantial correlation between TCGA (PRO, MES, DIF, IMR) and Tothill (C1, C2, C4, C5) subtypes. The MES and PRO TCGA subtypes demonstrated marker overlap with the C1 and C5 Tothill subtypes, while the majority of DIF and IMR cases were of the C4 and C2 subtypes, respectively. This overlap is consistent with previous reports of overlap between these subtyping approaches [19].

When comparing genomic features of these subtypes, the IMR/C2 groups demonstrated enrichment for *BRCA1*/*2*m. These cases also demonstrated the highest immune cell infiltration burden, with significantly greater levels of CD3+ and CD8+ cell infiltration. Together, the high *BRCA1*/*2*m rate and high levels of immune engagement in IMR/C2 tumours likely underpin the favourable outcome reported in these patient groups [19, 28]. In contrast, the vast majority of PRO/C5 cases were *BRCA1*/*2* wild-type, and instead demonstrated the highest rates of *CCNE1*g; the PRO/C5 subtypes may therefore represent the group least likely to benefit from PARP inhibition. Previous reports have suggested that PRO cases may derive greatest benefit from anti-angiogenic agents such as bevacizumab [22]; the low *BRCA1*/*2*m rate in this group may support the use of these agents over PARPi in this patient group. Conversely, the IMR/C2 group harbour a large number of *BRCA1*/*2*m patients that are likely to benefit from PARPi. Some investigators have suggested that anti-angiogenic therapies may not confer greatest benefit in patients with HGSOC demonstrating immune-related gene expression signatures [17], or may not benefit some groups of HRR-deficient patients [15, 30, 31]. Together, these data suggest that further dissection of the relationship between transcriptional subtypes, HRR status and relative benefit of single versus combined PARPi/anti-angiogenic strategies is required.

PRO/C5 cases were also uniformly low in tumour-infiltrating CD3+ and CD8+ cells, suggesting poor engagement of the immune system against the tumour within this patient group; this may partially account for the shorter survival time previously described in these cases [19, 28]. These data suggest that immune checkpoint inhibitors, currently under investigation in ovarian cancer, are also unlikely to represent viable therapeutic options for improving survival time in these patients [32].

Our multi-layer characterisation also sheds further light upon HRR pathway players and their importance in HGSOC. We show that *EMSY*-overexpressing cases appear *BRCA2*m-like in their survival profile and therapy sensitivity – consistent with EMSY’s role as a BRCA2 regulator [16]. However, they do not appear to be over-represented in the IMR/C2 transcriptomic subtypes, and do not demonstrate a higher burden of tumour-infiltrating immune cells. We also demonstrate the importance of aberrations in HRR genes regardless of the transcriptional subtype context; the hazard ratio for cases with HRR gene aberrations ranged between 0.48-0.68 across all transcriptional subtypes. These data confirm that the survival benefit among patients with tumours displaying HRR gene aberrations is not due to differential distribution of transcriptional subtypes.

*CCNE1*g has been the focus of intense research interest since its identification as a recurrent event in HGSOC [5, 33-36]. A number of studies have suggested that cases harbouring *CCNE1*g have poorer survival, with some suggesting this is due to greater intrinsic chemoresistance [5, 33-35]. However, these comparisons have typically been made against the wider non-*CCNE1*g population without accounting for HRR-deficiency, which is associated with longer survival and increased platinum sensitivity, confounding these comparisons. We compare the *CCNE1*g population directly to non-*CCNE1*g HHRwt cases. *CCNE1*g was not associated with significantly poorer response rate to first-line chemotherapy, or chemotherapy for relapsed disease, within our cohort. We show that, although the most advanced stage cases are under-represented in the *CCNE1*g group, *CCNE1*g cases demonstrate shorter survival time and that their survival is significantly poorer compared to non-*CCNE1*g HRRwt patients upon multivariable analysis. *CCNE1*g tumours also demonstrated the lowest levels of infiltrating immune cells compared to the other HRR-centric groups, which may contribute toward the shorter patient survival time. Mutual exclusivity of *CCNE1*g and HRR gene events suggests that the former are likely to represent a patient group who benefit least from PARP inhibition. Moreover, the low immune infiltration levels demonstrated by these cases suggests that immune checkpoint inhibitors are unlikely to be effective in these patients [32]. Given that *CCNE1*g is most frequent in the PRO/C5 transcriptomic subtypes [22], and that the PRO subtype has been associated with greatest benefit from bevacizumab in some reports, *CCNE1*g cases may represent those likely to derive benefit from anti-angiogenic therapies. *CCNE1*g cases – alongside other HRR-proficient patient groups – represent HGSOC patients with shorter survival time for which new treatment approaches are needed to improve survival. However, the low frequency of other molecular events in *CCNE1*g cases (*BRCA1*/*2* wild-type, RB intact, PTEN intact, low immune cell infiltration), represents a challenge toward identifying further candidate biologically-targeted strategies within this patient group. Inhibition of WEE1 kinase represents a potential strategy of interest for CCNE1g cases, with recent data demonstrating objective responses to WEE1 inhibitors in treatment-refractory CCNE1g HGSOC [37].

Disruption of *PTEN* and *RB1* have only recently been identified as highly recurrent events in HGSOC [6]. The relationship of these events to other molecular features and their impact on patient outcome is poorly understood. We demonstrate that PTEN and RB protein loss are neither mutually exclusive with one another, nor mutually exclusive with other recurrent genomic events in this tumour type. Indeed, the frequency of RB loss was significantly higher in HRR-deficient cases and there was significant co-occurrence between RB and PTEN loss. By contrast, RB loss was a rare event in *CCNE1*g cases. Cases with RB or PTEN loss demonstrated reduced CN at their respective loci; however, not all cases with loss demonstrated low CN, suggesting mechanisms of inactivation beyond CN loss, consistent with reports of complex structural variants (SVs) affecting both *RB1* and *PTEN* [6]. Perhaps most interestingly, RB status discriminated outcome within the cases showing HRR gene aberrations, with the RB loss significantly associated with longer survival; RB-loss did not significantly impact outcome in cases without identifiable events in HRR genes. It is unclear whether this phenotype is due to differences in therapy sensitivity, or whether concurrent RB loss results in HRR-aberrant tumours with more indolent behaviour. Mechanistic work investigating the phenotypic and signalling consequences of RB loss in the context of HRR-deficiency is now warranted, including investigation of the relative chemosensitivity of RB-lost and RB-intact HRR-deficient cells.

We present a large, pathologically-confirmed HGSOC patient cohort with extensive follow-up and detailed clinical annotation, including chemotherapy response data. Together with the multiple layers of molecular characterisation, these represent major strengths of this work. However, we were unable to characterise genome-wide SVs due to a lack of whole genome sequencing, which is a limitation of the study. Lack of *BRCA1/2* SV and *BRCA1* promoter methylation data will have likely resulted in a more conservative HR estimate when comparing our HRR-aberrant and HRRwt populations. SVs such as translocations and inversions are known to affect *NF1* in a proportion of HGSOC patients [6], and we were unable to characterise this patient group in our study. Future work should seek to provide even greater resolution within the HRR-proficient patient population, including characterisation of cases with *NF1* loss.

## 5. CONCLUSION

Together, these data provide a high resolution picture of the molecular landscape in HGSOC, integrating genomic sequencing with copy number data, transcriptomic profiling and immune cell infiltration burden in a cohort of HGSOC with rich clinical annotation. Specific transcriptomic subtypes are associated with marked differences in frequency of HRR gene aberrations, *CCNE1*g and infiltration of immune cells; integration of these data highlights patient groups most likely to responding to conventional chemotherapy and targeted biological therapeutics. Patients with *CCNE1*g and HRRwt tumours represent those with greatest unmet clinical need; investigations of new treatment strategies should focus on this patient group. RB and PTEN loss are common in HGSOC and frequently occur alongside other molecular events, with RB loss affecting a large number of tumours with HRR gene aberrations.

## Data Availability

All data produced in the present study are available upon reasonable request to the authors

## Authors’ contributions

RLH: conceptualisation, formal analysis, investigation, methodology, visualisation, writing – original draft. AMM: methodology, investigation, writing – review and editing. COM: data curation, resources, writing – review and editing. TR: data curation, investigation, writing – review and editing. MC: data curation, project administration, writing – review and editing. AHP: investigation, writing – review and editing. IC: investigation, writing – review and editing. WGM: investigation, writing – review and editing. ARWW: investigation, writing – review and editing. CB: data curation, writing – review and editing. YI: investigation, writing – review and editing. AO: supervision, writing – review and editing. BD: methodology, writing – review and editing. JCB: conceptualisation, supervision, writing – review and editing. RM: conceptualisation, supervision, writing – review and editing. AM: conceptualisation, supervision, writing – review and editing. PR: conceptualisation, supervision, writing – review and editing. CAS: methodology, supervision, writing – review and editing. DPH: methodology, writing – review and editing. RK: methodology, writing – review and editing. CSH: conceptualisation, investigation, methodology, supervision, writing – review and editing. CG: conceptualisation, methodology, funding acquisition, supervision, writing – review and editing.

## Acknowledgements

We thank the patients who contributed to this study and the Edinburgh Ovarian Cancer Database from which the clinical data reported here were retrieved. We are grateful to the NRS Lothian Human Annotated Bioresource, NHS Lothian Department of Pathology and Edinburgh Experimental Cancer Medicine Centre for their support. We thank the Edinburgh Clinical Research Facility, Western General Hospital, Edinburgh, UK for their support with the high throughput sequencing described here.

