## supplementary materials for "Multiomic characterisation of high grade serous ovarian carcinoma enables high resolution patient stratification"

**SUPPLEMENTARY INFORMATION**

**SUPPLEMENTARY METHODS**

***1. Pathology review and immunohistochemistry for WT1 and p53***

H&E-stained slides underwent pathology review by two expert gynaecological pathologists (ARWW, WGM) prior to the initial transcriptomic characterisation of these samples [1, 2]. Prior to inclusion in the matched genomic-transcriptomic HGSOC study, cases were subject to additional pathology review (CSH); cases uncertain to represent HGSOC (n=26) underwent IHC for WT1 and p53 to aid histotyping (HGSOC: WT1 positive, p53 aberrant expression pattern) (figure S1).

WT1 and p53 IHC was performed on the Leica BOND III Autostainer using IHC protocol F with 1:1000 anti-WT1 6F-H2 antibody (DAKO) or 1:50 anti-p53 DO-7 antibody (DAKO). For WT1, positive staining was defined as positive tumour nuclei; negative staining was defined as no tumour nuclear staining with corresponding positive stromal cells. For p53, aberrant positive diffuse tumour nuclear staining or complete absence of tumour nuclear staining was defined as aberrant expression [3]; variable nuclear intensity was defined as wild-type pattern. Stromal cells served as an internal positive control for both markers.

***2. CCNE1 and EMSY copy number assays***

*CCNE1* and *EMSY* copy number (CN) were quantified by TaqMan qPCR Copy Number Assays (Hs07158517_cn and Hs06316346_cn, ThermoFisher Scientific) using the StepOne Plus Real-Time PCR System (Applied Biosystems, ThermoFisher Scientific) and StepOne Software Version 2.3 with 10ng template DNA as determined by HS qubit assay. RNaseP reference assay was used as a copy number reference assay. NA12878 human reference DNA was purchased from the Coriell Institute and included in each run. CN variants were called with CopyCaller v2.0 software using NA12878 as a calibrator sample (CN=2).

*CCNE1* copy number gain (*CCNE1*g) was defined as ≥4 *CCNE1* copies. *EMSY* amplification was defined as ≥6 copies of *EMSY*. FUOV1 and OVCAR3 cell line DNA samples were included as controls gain of *CCNE1* and *EMSY*, respectively.

***3. Custom Integrated DNA Technologies Gene Capture Panel***

High throughput sequencing was performed using a custom Integrated DNA Technologies (IDT) gene capture panel with unique molecular indices (UMIs); the panel was designed to capture all exonic regions of: *ABCB1*, *AC004223.3*, *ARID1A*, *ATM*, *ATR*, *ATRX*, *BAP1*, *BARD1*, *BCL2L1*, *BLM*, *BRAF*, *BRCA1*, *BRCA2*, *BRIP1*, *C11orf65*, *CCNE1*, *CDK12*, *CHD4*, *CHEK1*, *CHEK2*, *CTNNB1*, *EGFR*, *EMSY*, *ERBB2*, *ERCC4*, *EZH2*, *FANCA*, *FANCB*, *FANCC*, *FANCD2*, *FANCE*, *FANCF*, *FANCG*, *FANCI*, *FANCL*, *FANCM*, *GNAS*, *KIT*, *KRAS*, *MAD2L2*, *MDM2*, *MLH1*, *MRE11*, *MSH2*, *MSH6*, *MUS81*, *MUTYH*, *NBN*, *NDUFB2*, *NF1*, *NF2*, *NRAS*, *PALB2*, *PARP1*, *PARP2*, *PAXIP1*, *PDGFRA*, *PER3*, *PIK3CA*, *PMS2*, *PPP2R1A*, *PPP2R2A*, *PRKDC*, *PTEN*, *RAD50*, *RAD51*, *RAD51B*, *RAD51C*, *RAD54L*, *RB1*, *RNASEH2A*, *RNASEH2B*, *RNASEH2C*, *RPA1*, *RUNDC3B*, *SHFM1*, *SLC25A40*, *SLFN11*, *SLX4*, *TOE1*, *TP53*, *TP53BP1*, *UBE2T*, *VRK2.* Whole genome libraries were generated using 200ng input DNA and pooled into groups of 16 for target gene capture and sequencing using an Illumina NextSeq 550 at the Edinburgh Clinical Research Facility, Western General Hospital, Edinburgh, UK. The median per-sample mean target coverage was 593X (range 205-3278X).

***4. Processing of sequencing data and variant calling***

Sequence reads were processed using the bcbio v1.0.6 high throughput sequence analysis pipeline (Supplementary methods section 4): reads were aligned to hg38 with bwa v0.7.17, sorted and duplicates marked with bamsormadup (biobambam v2.0.79), UMIs were added as tags with umis v0.9.0b0, files were converted to BAM format and indexed using samtools v1.6. Reads were then grouped by UMI, and consensus reads were called and filtered with fgbio v0.4.0.Consensus reads were extracted with bamtofastq (biobambam) and re-aligned, sorted and indexed. The aligned consensus reads underwent base quality score recalibration with the Genome Analysis Toolkit (GATK) v3.8 and variant calling was performed using a majority vote system from three variant callers (Freebayes v1.1.0.46 [4], VarDict Java v1.5.1 [5], and GATK Mutect2 [6]). The DKFZ bias filter was applied to identify likely false positive variants caused by strand bias or FFPE-induced DNA damage. Owing to the reported ubiquitous p53 disruption in HGSOC, *TP53*-wildtype cases underwent manual review of aligned reads in IGV to confirm wild-type status; 24 further mutations were identified by manual review, the vast majority of which (n=20) were splice site mutations toward read ends.

***5. Filtering of called variants***

Called variants at a minimum 10% allele frequency were annotated using the Ensembl VEP v90.9 against Ensembl release 90 and filtered using VEP annotation and the ClinVar database [7] to retain only likely functional variation: variants documented as pathogenic were retained as mutations, and those documented as benign were filtered. Within the remaining callset, nonsense mutations, frameshifting indels and splice site variants were retained as likely detrimental variants. Remaining synonymous, missense non-coding and undocumented significance variants were filtered as variants of uncertain significance.

***6. Transcriptomic characterisation and subtyping***

Gene expression data were generated as part of a previous study identifying transcriptionally-defined molecular subtypes of HGSOC [1, 2]. Samples were characterised in a larger training cohort (n=247 HGSOC in the present study), and a subsequent validation cohort (n=115 HGSOC in the present study). RNA was extracted from macrodissected FFPE tumor material using the Roche High Pure FFPE RNA Isolation kit, cDNA was amplified using the NuGEN FFPE WT-Ovation FFPE System kit, then fragmented and labelled using the NuGEN Encore Biotin Module. Resultant products were hybridisation to the Ovarian DSA™ cDNA microarray platform. Each cohort was pre-processed using the Robust Multi-Array Average (RMA) method prior to a quality control.

TCGA- and Tothill-based transcriptomic subtypes were determined using the ConsensusOv R package [8] with the 'ConsensusOv' and 'Helland' approaches.

*EMSY* overexpression was defined as expression within the top 14% of cases, as indicated recommended by the previous *EMSY* expression study [2] (status already available for the training cases from the previous study, and determined for the validation cases accordingly).

***7. Immune cell infiltration analysis***

Tumour infiltrating CD3-positive and CD8-positive immune cells were quantified by immunohistochemistry of tumour tissue microarrays (TMAs); three 0.8mm cores were taken from a tumour-containing FFPE tumour block per patient to construct the HGSOC cohort TMA. 4um TMA sections were stained for CD3 and CD8 using the Leica BOND III Autostainer and Leica BOND ready-to-use anti-CD3 and anti-CD8 antibodies with IHC protocol F. Stained sections were imaged and analysed using QuPath version 0.1.2. Tumour area was marked as a region of interest and positive and negative cells were counted using the positive cell detection protocol. Where cases were unevaluable due to damaged/missing cores (n=24 for CD8, n=24 for CD3), whole slide 4um FFPE sections were stained for CD3 and CD8 where available (n=21 for CD8, n=21 for CD3) and virtual TMAs were constructed using random sampling of 3 tumour-containing regions equivalent to the area of a TMA core. These were then processed as above.

Automated positive cell quantification was validated by manual scoring of a subset of tumour-containing cores by two human observers (RLH, AHP) (180 randomly selected cores per marker), demonstrating excellent correlation between human and machine scoring (spearman's rho>0.95, P<0.0001 for both observers against QuPath).

Positive infiltrating cell burden was quantified as the percentage of positive cells within tumour islets.

***8. Immunohistochemistry for PTEN and RB***

PTEN and RB protein loss was detected by IHC using sections of the HGSOC TMA. PTEN IHC used 1:50 M3627 clone 6H2.1 (DAKO); RB IHC used 1:100 NCL-L-RB-358 (Leica). Loss was defined as complete loss of positive staining in tumour cells with positive adjacent stromal staining. Wild-type pattern was defined as positive tumour cell staining. Two observers scored each core independently (RLH, YI). Disagreement was resolved by subsequent discussion to reach a consensus call; where a consensus was not agreed, staining was regarded as non-evaluable.

Where cases were unevaluable due to damaged/missing cores or equivocal staining (n=33 for PTEN, n=44 for RB), whole slide 4µm FFPE sections were stained for PTEN and RB where available (n=21 for PTEN, n=34 for RB) and scored as above.

***9.Copy number analysis from off-target sequencing reads***

Aligned bam files produced by the bcbio nextgen workflow were used for further CN analysis. Relative CN for 50kB segments of the genome were determine using the CopywriteR R package [9], whereby off-target reads are used to produce genome-wide CN estimates.

CN loss events were defined as regions with a log2 CN ratio of ≤ -2; CN gain events were defined as regions with a log2 CN ratio of ≥1.5. For estimating CN of *RB1* and *PTEN*, the mean CN across the 50kB segments encompassing *RB1* and *PTEN* were calculated as the overall gene CN. For quantification of total CN gains, adjacent 50kB segments that demonstrated CN gain were merged to be counted as a single large CN gain events (using a 10% tolerance for the CN gain threshold in adjacent segments). The same approach was applied when quantifying the total number of loss events.

*10. Response and progression data*

Radiological response to first- and second-line chemotherapy was defined using measured change in disease volume as calculated using the product of longest diameter and longest perpendicular diameter: complete response was defined as complete resolution of pre-treatment disease, partial response (PR) was defined as disease reduction by ≥50%, progressive disease (PD) was defined as radiologically-confirmed appearance of new lesions or ≥50% increase in tumour size (by bi-dimensional measurements). Evaluable cases not reaching criteria for PR or PD were classified as stable disease (SD).

CA125 tumour marker response was evaluated using GCIG criteria [10]: complete response (GCIG-CR); was defined as confirmed normalisation of CA125 after a pre-treatment baseline value at least twice the upper limit of normal; partial response (GCIG-50%) was defined as confirmed reduction of CA125 by at least 50% from a baseline value at least twice the upper limit of normal. CA125 progression was defined as confirmed doubling of CA125. Evaluable cases not reaching criteria for response or PD were classified as no change in CA125.

Progression-free survival (PFS) was defined as the time from pathologically confirmed diagnosis to first progression event (radiological PD, radiologically confirmed recurrence or CA125 progression by GCIG criteria). 32 cases were non-evaluable for PFS time due to insufficient investigations.

### SUPPLEMENTARY TABLES

Supplementary table 1. Identified mutations from targeted sequencing of 362 HGSOC cases

| Gene | HGSOC cases with mutation | % |
|---|---|---|
| *TP53* | 355 | 98.1 |
| *BRCA1* | 46 | 12.7 |
| *BRCA2* | 24 | 6.6 |
| *RB1* | 11 | 3.0 |
| *NF1* | 10 | 2.8 |
| *NF2, PIK3CA* | 8 | 2.2 |
| *CDK12* | 5 | 1.4 |
| *ARID1A* | 4 | 1.1 |
| *FANCA, KRAS, SLFN11, PER3, BRIP1* | 3 | 0.8 |
| *MSH6, CTNNB1, SLX4, CHEK2, PRKDC* | 2 | 0.6 |
| *CHD4, AC004223.3, PTEN, EMSY, FANCF, BRAF, PARP2, PAXIP1, ATM, CCNE1, RAD51C, BAP1, NBN, PALB2, FANCM, TP53BP1, GNAS, FANCC, RNASEH2B, PPP2R1A, MSH2, SLC25A40, ERCC4* | 1 | 0.3 |
| *ABCB1, ATR, ATRX, BARD1, BCL2L1, BLM, C11orf65, CHEK1, EGFR, ERBB2, EZH2, FANCB, FANCD2, FANCE, FANCG, FANCI, FANCL, KIT, MAD2L2, MDM2, MLH1, MRE11, MUS81, MUTYH, NDUFB2, NRAS, PARP1, PDGFRA, PMS2, PPP2R2A, RAD50, RAD51, RAD51B, RAD54L, RNASEH2A, RNASEH2C, RPA1, RUNDC3B, SHFM1, TOE1, UBE2T, VRK2* | 0 | 0.0 |

HGSOC, high grade serous ovarian carcinoma

Supplementary table 2. Comparison of *EMSY* overexpression versus copy number status

| | ***EMSY* expression status** | |
|---|---|---|
| | Overexpressed | Wild-type |
| ***EMSY* CN status** | | |
| Amplified | 10 | 14 |
| Non-amplified | 42 | 296 |
| Chi-squared test P<0.001 | | |

For amplification status as a predictor of overexpression: positive predictive value 0.42 (95% CI 0.22-0.63); negative predictive value 0.88 (95% CI 0.84-0.91); sensitivity 0.19 (95% CI 0.10-0.33); specificity 0.95 (95% CI 0.93-0.98)

### SUPPLEMENTARY FIGURES

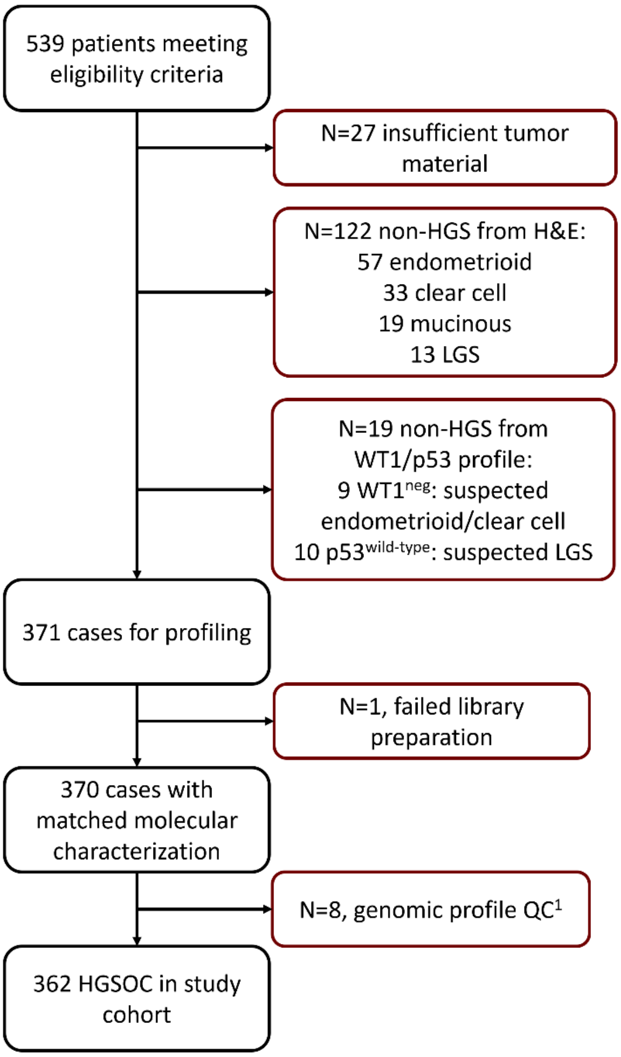


Figure S1. Case flow diagram for high grade serous ovarian carcinoma (HGSOC) cohort. [1]Excluded as likely non-HGS from genomic profile: *TP53* wild-type with mutation of *ARID1A*, *KRAS*, *PIK3CA* or *CTNNB1*. QC, quality control. LGS, low grade serous.

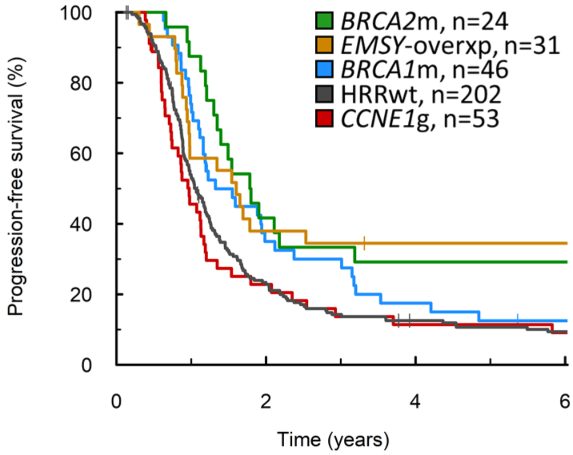


Figure S2. Progression-free survival of homologous recombination repair (HRR)-centric subtypes. *BRCA2*m, *BRCA2* mutant; *BRCA1*m, *BRCA1* mutant; *EMSY*-overxp; overexpression of *EMSY*; *CCNE1*g, gain of *CCNE1*; HRRwt, non-*CCNE1*g HRR wild-type.

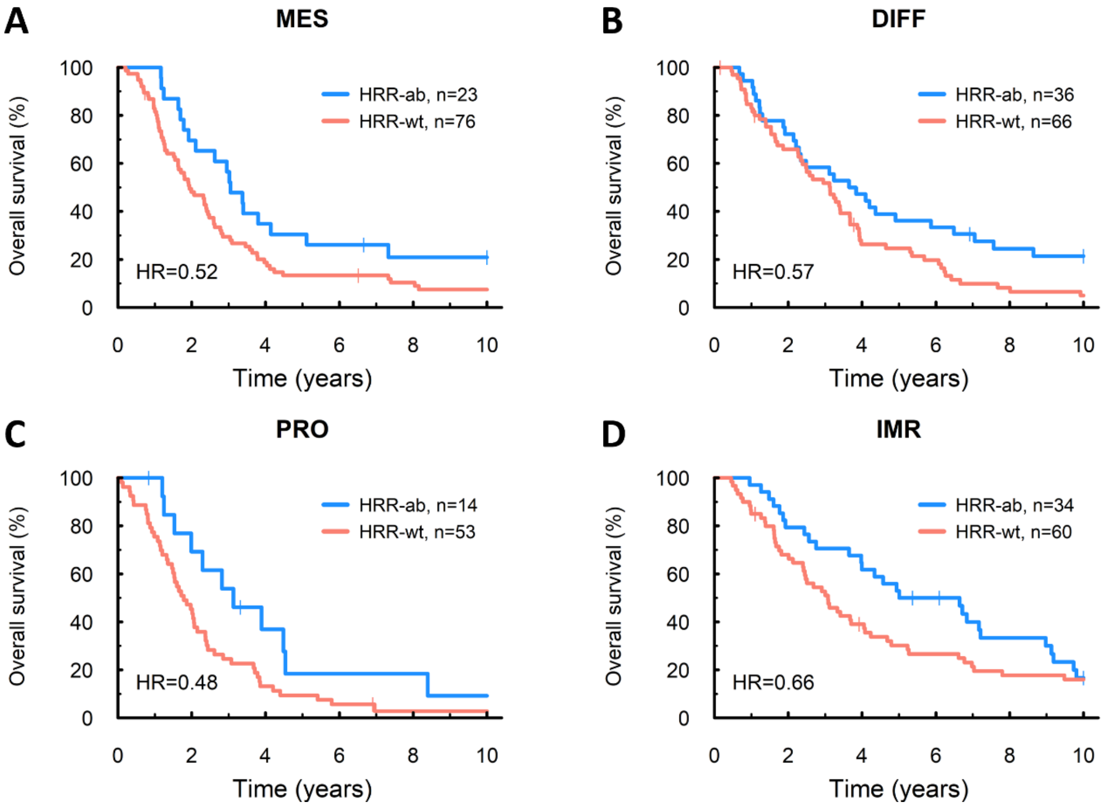


Figure S3. Impact of homologous recombination repair aberrations (HRR-aberrant: *BRCA1* mutation, *BRCA2* mutation, *EMSY*-overexpression or non-*BRCA* HRR gene mutation) on overall survival within TCGA transcriptomic subtypes. (A) Overall survival within the MES subtype. (B) Overall survival within the DIFF subtype. (C) Overall survival within the PRO subtype. (D) Overall survival within the IMR subtype. HRR-ab, HRR-aberrant; HRR-wt, HRR wild-type reference population: *CCNE1*-gained plus other HRR wild-type cases.

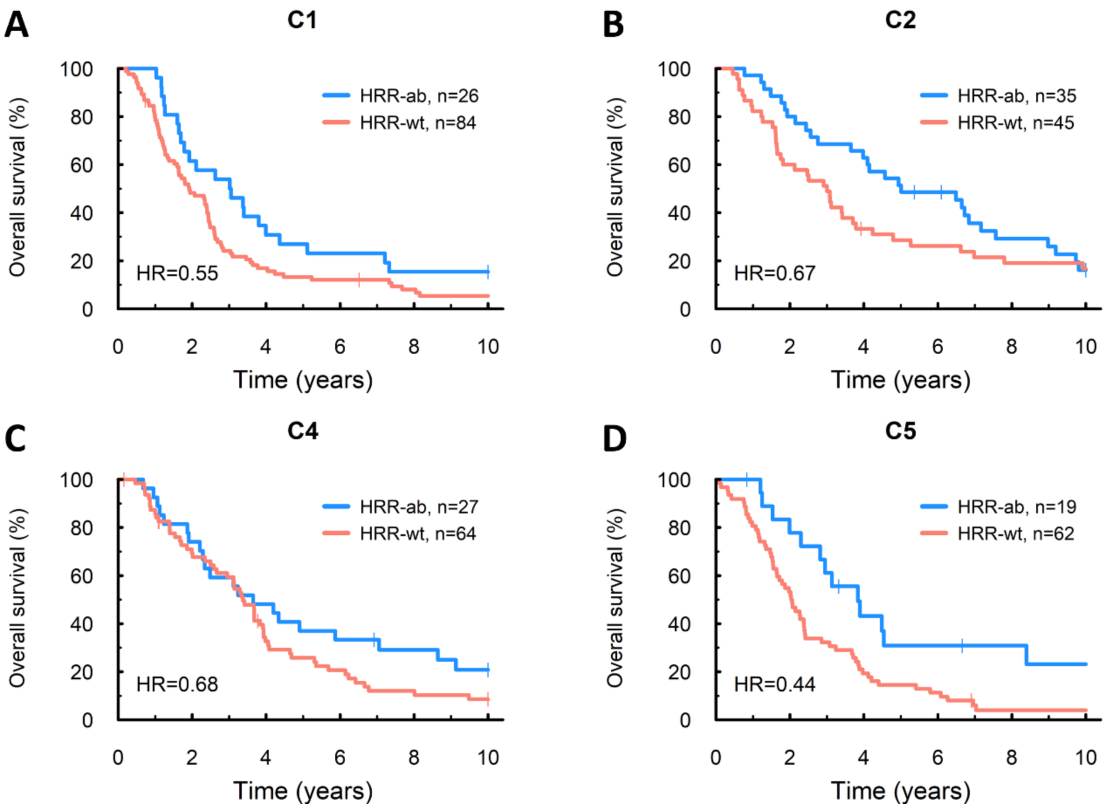


Figure S4. Impact of homologous recombination repair aberrations (HRR-aberrant: *BRCA1* mutation, *BRCA2* mutation, *EMSY*-overexpression or non-*BRCA* HRR gene mutation) on overall survival within Tothill transcriptomic subtypes. (A) Overall survival within the C1 subtype. (B) Overall survival within the C2 subtype. (C) Overall survival within the C4 subtype. (D) Overall survival within the C5 subtype. HRR-ab, HRR-aberrant; HRR-wt, HRR wild-type reference population: *CCNE1*-gained plus other HRR wild-type cases.

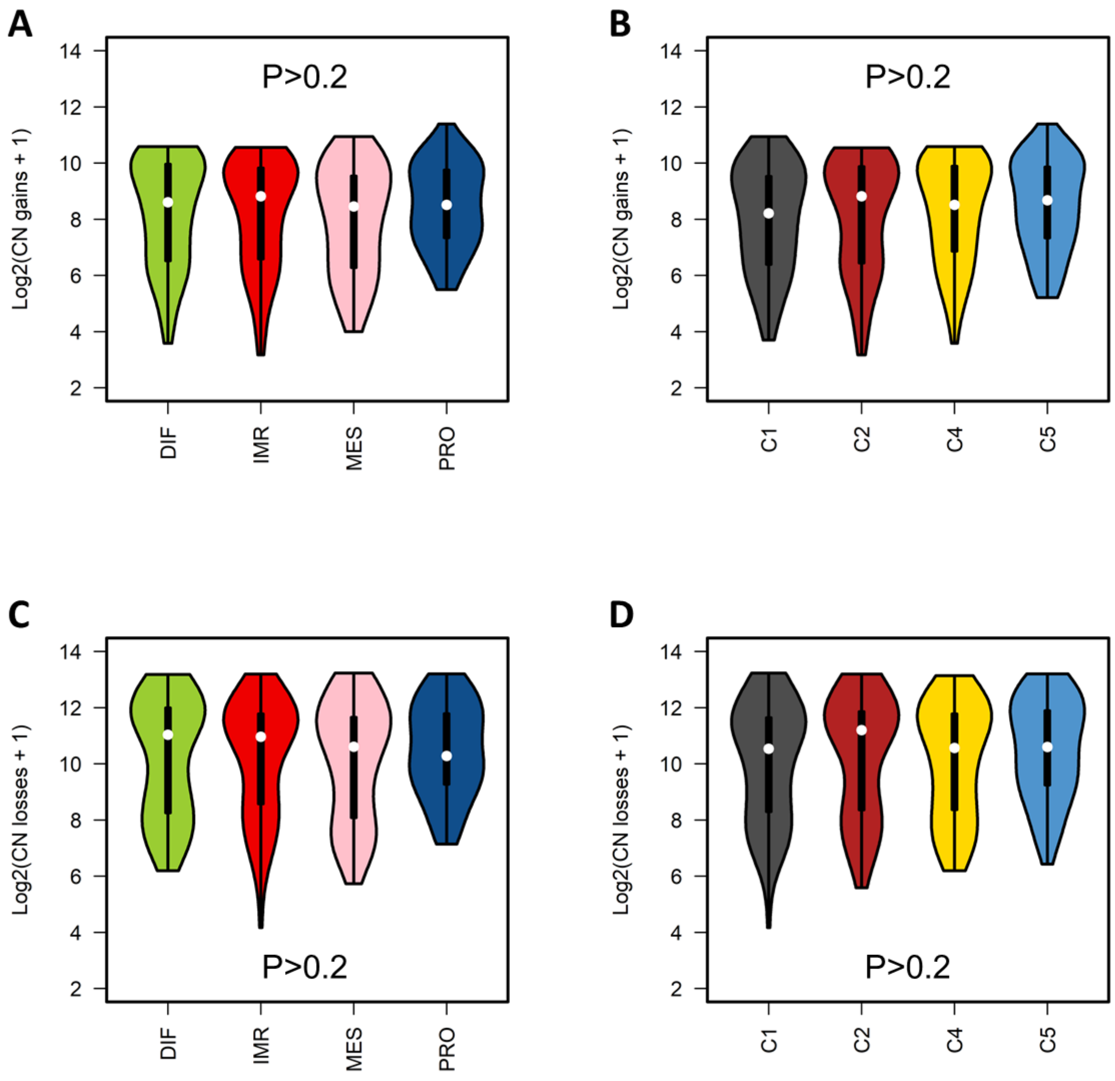


Figure S5. Violin plots of copy number (CN) gain and CN loss event burden across transcriptomic subtypes of high grade serous ovarian carcinoma. (A) CN gain event burden across TCGA transcriptomic subtypes. (B) CN gain event burden across Tothill transcriptomic subtypes. (C) CN loss event burden across TCGA transcriptomic subtypes. (D) CN loss event burden across Tothill transcriptomic subtypes.

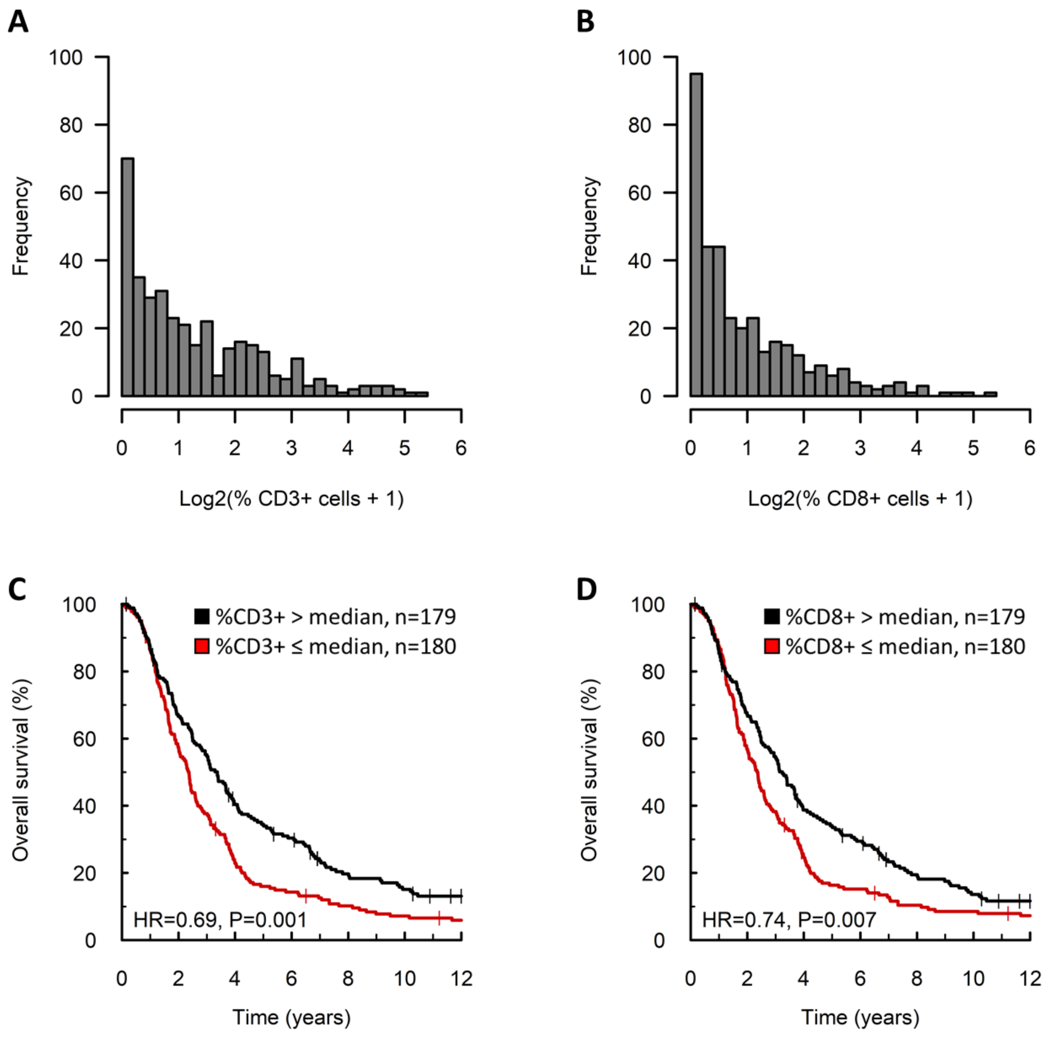


Figure S6. Tumour-infiltrating immune cells in high grade serous ovarian carcinoma. (A) Distribution of CD3+ infiltrating cell burden. (B) Distribution of CD8+ infiltrating cell burden. (C) Impact of CD3+ infiltrating cell burden on overall survival. (D) Impact of CD8+ infiltrating cell burden on overall survival.

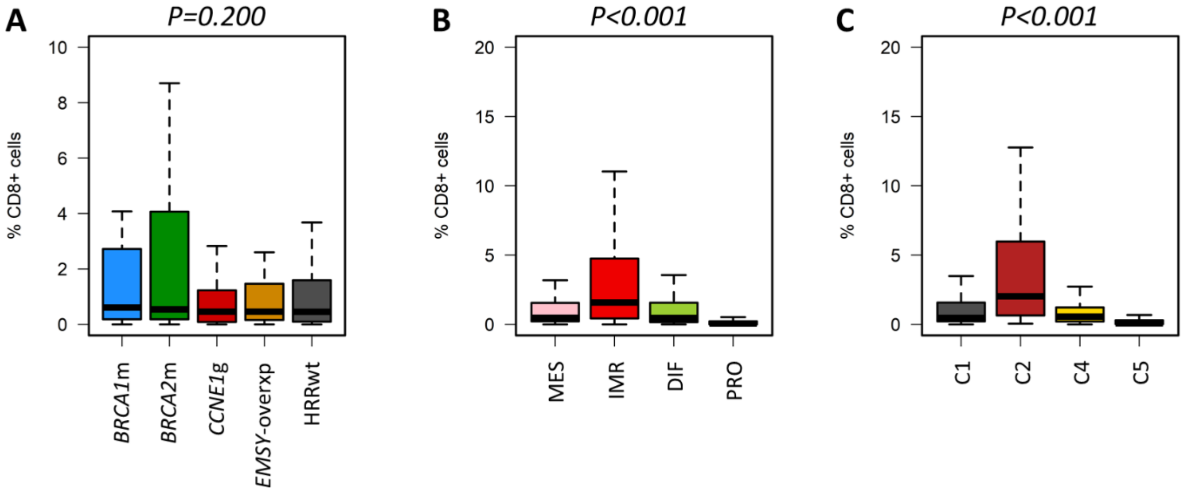


Figure S7. Tumour-infiltrating CD8+ cells across high grade serous carcinoma subtypes. (A) CD8+ infiltration across HRR-centric subtypes; labelled P value represents comparison of *BRCA2*m and *CCNE1*g groups using the Mann Whitney-U test. (B) CD8+ infiltration across TCGA transcriptomic subtypes; labelled P value represents comparison of IMR and PRO groups using the Mann Whitney-U test. (C) CD8+ infiltration across Tothill transcriptomic subtypes; labelled P value represents comparison of C2 and C5 groups using the Mann Whitney-U test. *BRCA2*m, *BRCA2* mutant; *BRCA1*m, *BRCA1* mutant; *EMSY*-overxp; overexpression of *EMSY*; *CCNE1*g, gain of *CCNE1*; HRRwt, non-*CCNE1*g homologous recombination proficient.

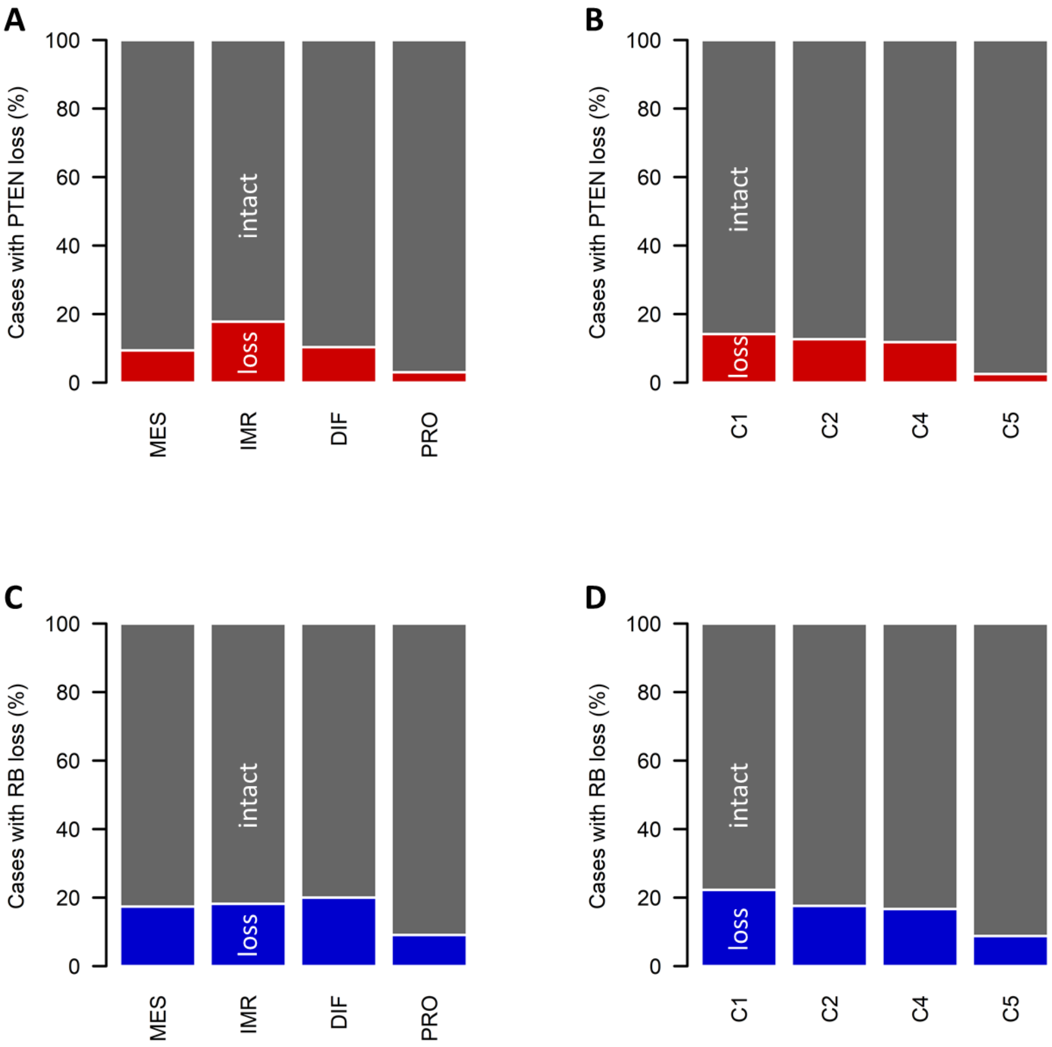


Figure S8. Loss of PTEN and RB protein expression across transcriptional subtypes of high grade serous carcinoma. (A) PTEN loss across TCGA transcriptomic subtypes. (B) PTEN loss across Tothill transcriptomic subtypes. (C) RB loss across TCGA transcriptomic subtypes. (D) RB loss across Tothill transcriptomic subtypes.

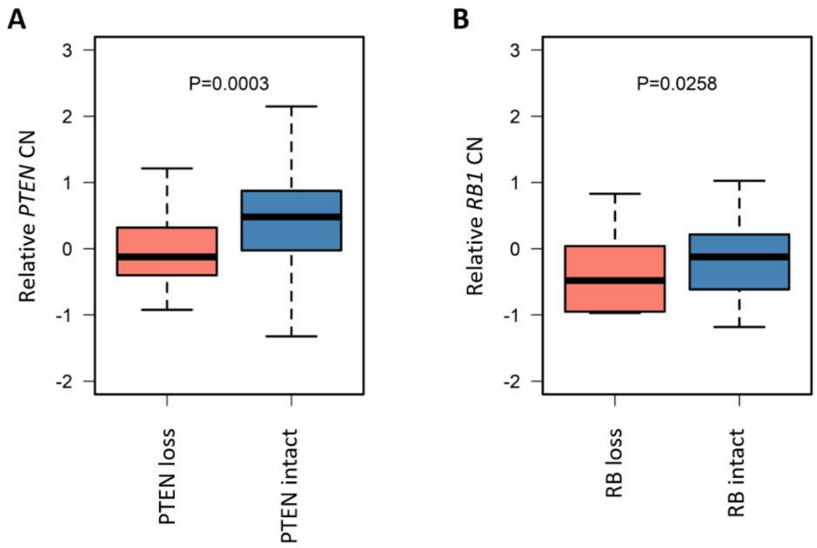


Figure S9. Calculated copy number (CN) of *PTEN* and *RB1* genes in cases with loss of PTEN and loss of RB expression, as determined by CopywriteR. (A) *PTEN* CN estimates between PTEN-lost and PTEN-intact cases. (B) *RB1* CN estimates between RB-lost and RB-intact cases.
